# A Framework for Interpreting White Blood Cell Counts From Lumbar Punctures in Pediatric Acute Lymphoblastic Leukemia

**DOI:** 10.1101/2025.09.02.25334951

**Authors:** Adith S. Arun

## Abstract

In pediatric acute lymphoblastic leukemia (ALL), central nervous system (CNS) involvement is staged using white blood cell (WBC) counts from lumbar punctures (LPs), microscopy of LP-derived cells, and CNS imaging. CNS staging informs the need for additional intrathecal chemotherapy, which can result in side-effects including significant neurotoxicity. However, nearly 20% of LPs are traumatic, or contaminated by peripheral blood. In these cases, the Steinherz-Bleyer (S-B) algorithm is used instead of WBC counts to determine CNS involvement. The intuition for this algorithm has not been presented. In this work, we conceptualize LP lab values as a mixture of blood and cerebrospinal fluid (CSF) and present this mixture as a convex combination problem. Then, we demonstrate that the S-B algorithm asks whether the CSF to blood WBC ratio is at or above that of the mixing ratio (i.e., contaminated to uncontaminated ratio). Additionally, we derive an expression for estimating the true but unobserved CSF WBC in traumatic LP cases such that the existing atraumtic CSF WBC guidelines may be used. Finally, we present a Bayesian approach to incorporate non-zero CSF red blood cell (RBC) counts and suggest that this biologically-motivated assumption underlying S-B is likely not clinically relevant for the majority of patients.

## Introduction

Pediatric acute lymphoblastic leukemia (ALL) treatment protocols differ based on the extent of central nervous system (CNS) involvement [1]. CNS involvement is determined by imaging and lumbar puncture (LP) and graded as either CNS-1, CNS-2, or CNS-3. LPs measure cerebrospinal fluid (CSF) white blood cell (WBC) counts and the presence of lymphoblasts (blasts) on microscopy. Absence of blasts and CSF WBC *<* 5 indicates CNS-1, presence of blasts and CSF WBC *<* 5 indicates CNS-2, and presence of blasts and CSF WBC *≥* 5 indicates CNS-3 [1, 2]. Patients are typically assigned CNS-3 status, regardless of CSF findings, if CNS imaging is consistent with leukemia or if neurological symptoms are present, most commonly cranial nerve palsies [3]. Given the high rates of CNS relapse, patients with ALL are given intrathecal therapy as CNS prophylaxis [4]. However, patients classified as CNS-3 receive additional intrathecal chemotherapy and may undergo craniospinal radiation, both of which are associated with short- and long-term neurotoxicities [1].

These criteria for CNS staging based on CSF results are established for LPs that are atraumatic, where only CSF is collected and no peripheral blood is drawn. Traumatic LPs occur when peripheral blood contaminates the CSF and are typically defined by a CSF red blood cell (RBC) count *≥* 10 per *µ*liter [1]. Traumatic LPs are estimated to have an incidence of about 20% in pediatric oncology [5]. Given the contamination of CSF with peripheral blood, the lab readout of the traumatic LP sample reports the WBC count of the contaminated sample. It does not report the true CSF WBC value. Therefore, the Steinherz-Bleyer (S-B) algorithm is often used to adjust for contamination from blood when determining CNS-3 status [1, 6, 7, 8].

The S-B algorithm classifies pediatric ALL patients with traumatic LPs as CNS-3 if the measured CSF WBC to RBC ratio is greater than or equal to two times the peripheral blood WBC to RBC ratio [2]. Otherwise, the patient is classified as CNS-2. To date, there does not exist work unpacking the assumptions behind S-B algorithm or explaining the mathematical intuition behind the formula. In this paper, we introduce a framework for conceptualizing lab values from LPs as a mixture of peripheral blood and CSF. We then provide the rationale for requiring the LP ratio to be at least twice the peripheral ratio in the S-B algorithm. Finally, we remove the core assumption behind the S-B algorithm and introduce a simple Bayesian method for estimating the true but unobserved WBC CSF count in traumatic LPs.

## Approach

First, we introduce a framework for describing LP lab results as a mixture of peripheral blood and CSF values. An atraumatic LP is a direct measurement of CSF WBC and RBC values. But, the WBC and RBC values measured in a traumatic LP, by definition, are a mixture of the values that would be found in peripheral blood and CSF. This situation can be expressed as a convex combination problem.

Let *b*_*r*_ and *b*_*w*_ be the measured RBC and WBC count per *µ*liter, respectively, from a patient’s blood. Similarly, let *z*_*r*_ and *z*_*w*_ be the measured RBC and WBC count per *µ*liter from a patient’s LP. Let *y*_*r*_ and *y*_*w*_ be the true but unobserved RBC and WBC count from the patient’s CSF. Let *p* be the fraction of blood in the LP and can take on any value from zero to one inclusive.

Therefore, we have

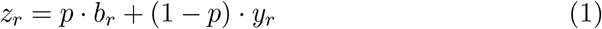

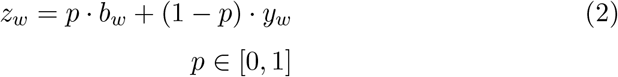

We observe that if the LP was completely atraumtic, then *p* = 0 and Equations 1 and 2 reduce down to *z*_*r*_ = *y*_*r*_ and *z*_*w*_ = *y*_*w*_, respectively. Thus, we measure the true CSF RBC and WBC count, as expected. When *p* = 1, and the LP is completely traumatic and the entire LP is just peripheral blood, *z*_*r*_ = *b*_*r*_ and *z*_*w*_ = *b*_*w*_.

### Derivation of rationale behind S-B algorithm

The S-B algorithm can be expressed as

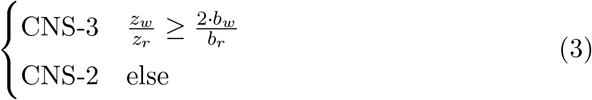

We can rearrange the S-B inequality and substitute Equations 1 and 2 for *z*_*r*_ and *z*_*w*_ to arrive at

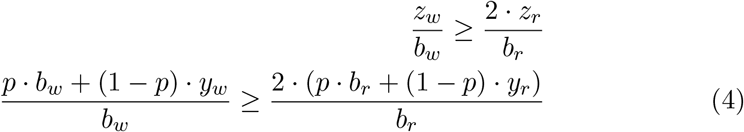

The assumption of the S-B algorithm is that the true but unobserved CSF RBC count is zero. This condition is generally true for patients without intracranial bleeding. Pediatric patients presenting exclusively with ALL should have a true CSF RBC count of zero [1, 2]. Thus we assume *y*_*r*_ = 0. This simplifies the right hand side of our inequality down to

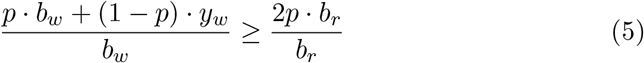

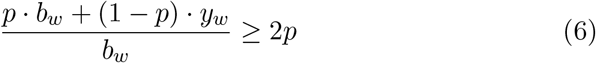

Then, we can rearrange the inequality as such

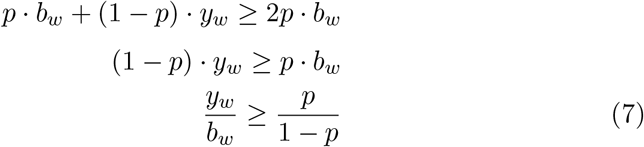

Note that since *p ∈* [0, 1], the expression 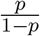 describes the odds ratio. In this situation, this odds ratio captures the relative contamination of blood in the LP. For example, if *p* = 0.2, the odds ratio is 0.25, which means that there is 1 part blood to 4 parts CSF in the LP. Thus, the S-B algorithm asks whether the ratio of true CSF WBC to blood WBC is at or above the mixing ratio.

### Estimation of CSF WBC counts in traumatic LPs

The S-B algorithm classifies the LP results as CNS-2 or CNS-3 but does not estimate *y*_*w*_, the true but unobserved count of WBC of the CSF. Estimation of *y*_*w*_ would allow for a numerical measure instead of a binary output. Also, with an estimate for *y*_*w*_, we can use the traditional atraumatic LP decision rules (i.e., CSF WBC *<* 5 or *≥* 5) for CNS classification instead of the S-B algorithm.

We can derive an expression for *y*_*w*_ using the same assumption as S-B: *y*_*r*_ = 0. First, rearrange Equation 1 to arrive at an expression for *p*

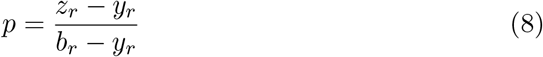

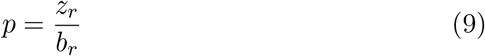

Then, we can substitute Eq. 9 for *p* in Eq. 2 and solve for *y*_*w*_.

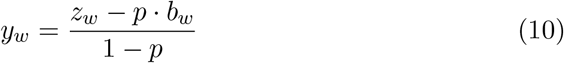

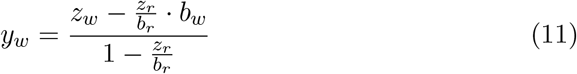

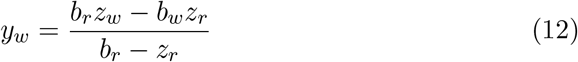

Thus, Equation 12 is a simple formula for estimating *y*_*w*_. But, just like S-B, it assumes *y*_*r*_ = 0. Though this assumption is biologically motivated and clinically reasonable, we seek to remove this constraint. To that end, we define a prior for *y*_*r*_. Namely, we define *θ* as the fraction of *z*_*r*_ truly found in *y*_*r*_. We have

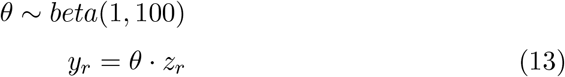

This prior reflects our clinical knowledge that most CSF RBC counts should be zero or near zero. This beta distribution has most of its density at zero with a right tail that decays on the interval from zero to one. Under this prior, the median value of *y*_*r*_ is 1% of *z*_*r*_.

We can draw Monte Carlo samples from *θ* and estimate *y*_*w*_ using Eq. 8 and Eq. 10. This yields a distribution of *y*_*w*_ values from which we can estimate the mean, median, and empirical confidence intervals for *y*_*w*_ as well as the fraction of Monte Carlo samples for which the S-B algorithm (using Eq. 7) assigns CNS-3 status. Using this approach, we allow *y*_*r*_≠ 0 and remove the primary assumption in S-B and Equation 12.

Code to run this numerical estimation strategy is publicly available at https://github.com/aditharun/lp-wbc-estimation.

## Results

We will work through a representative example taken from the American Society of Pediatric Hematology and Oncology and demonstrate the performance of S-B, naive estimation of *y*_*w*_ by assuming *y*_*r*_ = 0, and the Monte Carlo approach for estimating *y*_*w*_ [9]. Consider a patient with LP CSF WBC of 60 per *µ*liter, LP CSF RBC of 1500 per *µ*liter, blood WBC of 46000 per *µ*liter, and blood RBC of 3 *×* 10^6^ per *µ*liter.

The S-B (Eq. 3) inequality holds true (0.04 *≥* 0.03), and therefore the patient has CNS-3 disease. Next, we naively estimate *y*_*w*_ from Eq. 12 to be 37 per *µ*liter.

According to the CNS staging guidelines, since the estimated *y*_*w*_ *≥* 5, it suggests CNS-3 disease. The naive estimation of *y*_*w*_ is concordant with S-B. The fraction of blood in the LP is estimated to be 0.05% using Eq. 9.

We then apply the Monte Carlo approach to estimate *y*_*w*_. We raw 10,000 random samples from Eq. 13, estimate each *y*_*r*_, and compute the resulting *p* using Eq. 8 and *y*_*w*_ using Eq. 10 (Fig. 1).

**Figure 1:**
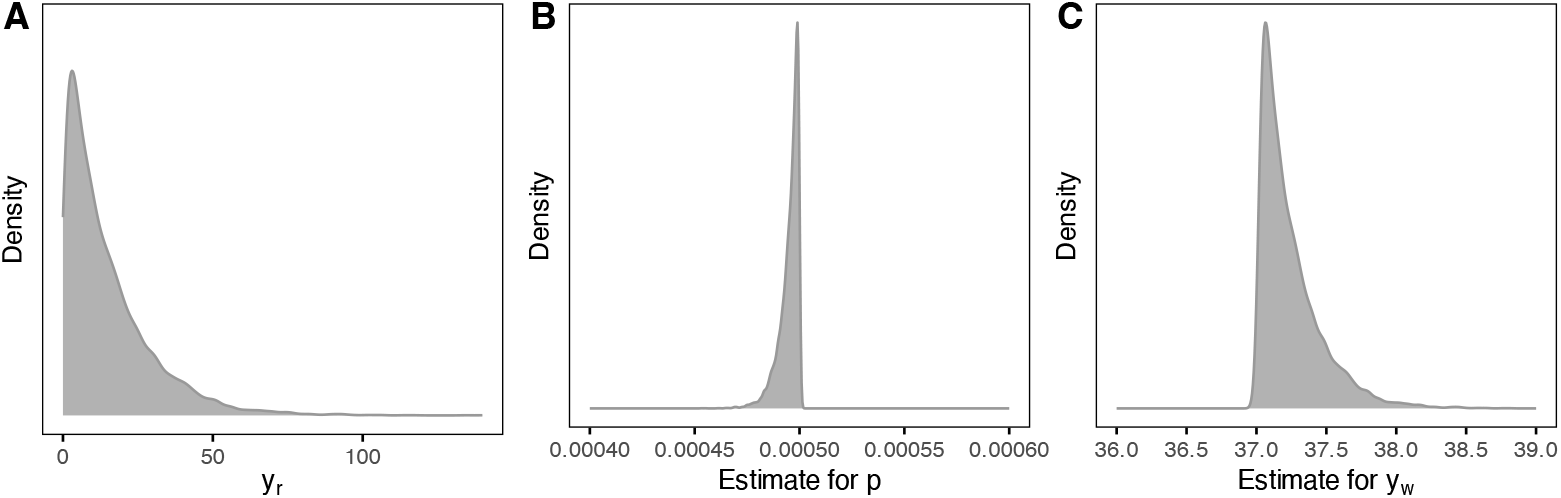
Sampled distribution for *y*_*r*_ (A) and posterior distributions of *p* (B) and *y*_*w*_ **(C)**. We consider an example with *z*_*r*_ = 1500, *z*_*w*_ = 60, *b*_*r*_ = 3 *×* 10^6^, and *b*_*w*_ = 46000 and incorporate our uncertainty in *y*_*r*_ to estimate *p* and *y*_*w*_ using Eqs. 10 and 8. The mean estimates for *y*_*r*_, *y*_*w*_ and *p* are 14.8 per *µ*liter, 37.2 per *µ*liter, and 0.05%, respectively.

The mean and 95% empirical confidence intervals for *y*_*r*_, *p*, and *y*_*w*_ are 14.8 [0.38 - 52.3], 0.0497% [0.0483% - 0.0499%], 37.2 [37.0 - 37.8] respectively (Fig. 1). The mean *y*_*w*_ when *y*_*r*_ *>* 0 is the same as *y*_*r*_ = 0. Also, we used the estimated *y*_*w*_ in each of the 10,000 samples and observed that all of the samples assigned CNS-3 status using S-B (Eq. 7). These results are concordant with the results when we assume *y*_*r*_ = 0.

The prior on *θ* is set such that most of the probability density lies at or near zero. We can relax this prior on *θ* to understand the sensitivity of *y*_*w*_ to our choice of *θ*. To accomplish this, we choose the biologically unlikely *uniform*(0, 0.5) with median value of *y*_*r*_ set to 25% of *z*_*r*_. Under this new prior, the median *y*_*w*_ estimate is 43 WBCs per *µ*liter. The relaxed prior generates a WBC estimate 14% higher than the biologically informed prior which does not impact clinical decision making in this scenario.

## Discussion

We introduce a framework for describing lab values from lumbar punctures as a convex combination of peripheral blood and true but unobserved CSF values. Using this approach, we show that the Steinherz-Bleyer (S-B) algorithm for staging CNS involvement in pediatric ALL patients assigns CNS-3 status only if the ratio of true CSF WBC to blood WBC is at or above the mixing ratio (i.e., the ratio of blood contaminating the sample to uncontaminated CSF fluid in the LP). Next, we introduce an expression to estimate the true CSF WBC count in a traumatic LP (Eq. 12). Direct estimation of CSF WBC count enables the use of atraumatic LP CNS staging guidelines [1, 2]. This method provides greater granularity than the binary threshold of the S-B algorithm.

The central assumption underlying S-B and our naive estimation of true CSF WBC counts in traumatic LPs (Eq. 12) is that true CSF RBC counts should be zero in atraumatic LPs in pediatric patients with no other pathologies except for ALL [2, 1]. We relax this clinically-informed assumption by allowing for non-zero CSF RBC counts and provide code for implementing this method. By incorporating uncertainty in CSF RBC counts, we can provide confidence intervals for estimated true CSF WBC and the fraction of Monte Carlo samples for which the S-B assigns CNS-3 status. These analyses resulted in narrow 95% confidence intervals with no change in CNS status assignment in the worked example. Allowing true CSF RBC to be greater than zero modestly increases the estimate for true CSF WBC. Using S-B will impact decision making only when CSF WBC counts are very low and near the staging guideline cutoff of 5 per *µ*liter. When true CSF WBC counts are at this decision boundary, the S-B algorithm has limited utility for clinical decision making. Other variables such as CNS imaging should guide treatment decisions in these situations. In practice, the S-B algorithm and equation for estimating true CSF WBC (Eq. 12) are likely valid for the vast majority of cases.

## Data Availability

All data produced in the present work are contained in the manuscript and code is available online at https://github.com/aditharun/lp-wbc-estimation

## Acknowledgments

We thank Dr. Gustavo Stolovitzky for his recommendations on this project.

